# Intergenerational and life-course links between a cardiovascular health-related methylation score and early vascular changes

**DOI:** 10.64898/2026.08.07.26359994

**Authors:** Binisha H. Mishra, Emma Raitoharju, Leo-Pekka Lyytikäinen, Nina Mononen, Juhani S. Koskinen, Jorma Viikari, Katja Pahkala, Suvi Rovio, Juha Mykkänen, Markus Juonala, Mika Kähönen, Olli T. Raitakari, Terho Lehtimäki, Pashupati P. Mishra

**Affiliations:** Department of Clinical Chemistry, Faculty of Medicine and Health Technology, Tampere University, Tampere, Finland; Finnish Cardiovascular Research Center Tampere, Faculty of Medicine and Health Technology, Tampere University, Tampere, Finland; Fimlab Laboratories, Tampere, Finland; Tampere University Hospital, Wellbeing services county of Pirkanmaa; Molecular Epidemiology Group, Faculty of Medicine and Health Technology, Tampere University, Tampere, Finland; Research Centre of Applied and Preventive Cardiovascular Medicine, University of Turku, Turku, Finland; Centre for Population Health Research, University of Turku and Turku University Hospital, Turku, Finland; Department of Medicine, University of Turku, Turku, Finland; Division of Medicine, Turku University Hospital, Turku, Finland; Department of Medicine, Satakunta Wellbeing Services County of Satakunta, Pori, Finland; Paavo Nurmi Centre & Unit for Health and Physical Activity, University of Turku, Turku, Finland; Department of Public Health, University of Turku and Turku University Hospital, Turku, Finland; Department of Clinical Physiology, Tampere University Hospital, Tampere Finland; Department of Clinical Physiology and Nuclear Medicine, Turku University Hospital, Turku, Finland; InFLAMES Research Flagship, University of Turku, Turku, Finland

## Abstract

**Background:** DNA methylation (DNAm) may capture cumulative genetic, environmental, and lifestyle influences on cardiovascular health. Composite DNAm score based on the American Heart Association Life’s Essential 8 (LE8) framework have been linked to clinical events, but their association with early vascular changes and intergenerational effects is unclear.

**Methods:** We studied up to 1432 participants from the multigenerational Young Finns Study (YFS-3G), including parents (G0) and adult offspring (G1). DNAm was measured using Illumina EPIC arrays in 2011 and/or 2018, and carotid intima–media thickness (cIMT) was assessed in 2018. The LE8 DNAm score was calculated as a weighted sum of methylation levels. Associations with cIMT were evaluated in intergenerational, prospective, and cross-sectional settings, adjusting for demographic, technical, and biological covariates and conventional cardiovascular risk factors.

**Results:** Higher parental LE8 DNAm score was associated with lower offspring cIMT (β = −0.022 mm/SD; p-value = 0.02), although the association was attenuated after adjustment for parental cardiovascular risk factors. In G1, a higher baseline DNAm score was associated with lower cIMT measured seven years later (β = −0.030 mm/SD; p-value = 1.1 × 10⁻⁵). This association remained significant after adjustment for follow-up cardiovascular risk factors (p-value=0.009) but not after additional adjustment for prior cIMT. Cross-sectionally, higher DNAm score was associated with lower cIMT in both generations, with attenuation after risk factor adjustment in G1 but not G0. Associations with carotid plaque were not significant. Genes associated with the DNAm score were enriched for immune and inflammatory pathways.

**Conclusions:** An LE8-derived DNAm score was associated with lower cIMT across the life course and, to a lesser extent, across generations. These findings suggest that blood DNAm reflects cumulative cardiovascular health and vascular burden and may complement conventional cardiovascular risk assessment.

## Background

Cardiovascular disease (CVD) remains the leading cause of morbidity and mortality worldwide, despite substantial advances in prevention and treatment. Atherosclerosis, the underlying pathology of most cardiovascular events, develops over decades and is influenced by a complex interplay of genetic, environmental, and lifestyle factors [WHO, 2025]. Traditional risk factors such as hypertension, dyslipidemia, smoking, and diabetes form the basis of widely used cardiovascular risk prediction tools, including the Framingham Risk Score [D’Agostino et al., 2008], and remain central to contemporary models such as QRISK3 [Hippisley-Cox et al., 2017] and SCORE2 [SCORE2 Working Group]. However, these risk factors do not fully capture individual variability in disease risk, particularly in the early, subclinical stages of vascular dysfunction [Gatto et al., 2020; Park et al., 2023; Barkas et al., 2024; Raitakari et al., 2025].

In recent years, epigenetic mechanisms, particularly DNA methylation (DNAm), have emerged as promising biomarkers linking environmental exposures to cardiovascular health (CVH) [Damiani et al., 2025]. DNAm reflects both genetic predisposition and cumulative environmental influences, including diet, physical activity, smoking, and psychosocial stress [Martin et al., 2018]. Epigenome-wide association studies (EWAS) have identified numerous CpG sites associated with cardiovascular risk factors and outcomes, suggesting that DNAm may provide integrative insight into disease processes that are not fully captured by traditional clinical measures [Aslibekyan et al., 2018; Westerman et al., 2020; Navas-Acien et al., 2021].

Building on this evidence, composite DNAm-based scores have been developed to summarize the epigenetic correlates of CVH [Zheng et al., 2022; Carbonneau et al., 2025]. Notably, DNAm signatures derived from the American Heart Association’s Life’s Essential 8 (LE8) framework incorporate information across multiple domains, including lipid levels, blood pressure, glycemic status, body composition, and health behaviors. These scores have been associated with cardiovascular risk and mortality, indicating their potential utility as holistic biomarkers of CVH [Carbonneau et al., 2025].

Despite these advances, several important gaps remain. First, most studies have focused on cross-sectional associations or clinical endpoints, with relatively limited investigation of subclinical vascular phenotypes, such as carotid intima-media thickness (cIMT), which reflect early atherosclerotic changes [Portilla-Fernández et al., 2021]. Second, while recent studies have begun to demonstrate the prospective utility of DNAm-based CVH scores, their broader longitudinal and life-course relevance is not well established [Zheng et al., 2022]. Understanding whether these scores predict future vascular changes would provide important evidence for their role in early disease development. Third, little is known about the intergenerational dimension of DNAm in CVH [Benincasa et al., 2024]. While familial clustering of cardiovascular risk is well documented, the extent to which epigenetic profiles in one generation are associated with vascular health in the next remains unclear.

The concept of intergenerational epigenetic influence is biologically plausible. Although most epigenetic marks are reprogrammed during development, growing evidence suggests that parental exposures and health status may influence offspring phenotype through both direct and indirect mechanisms, including shared environment, behavioral transmission, and potentially persistent epigenetic modifications [Fitz-James et al., 2022; Klibaner-Schiff et al., 2024]. Investigating these relationships in well-characterized multigenerational cohorts provides a unique opportunity to disentangle these pathways.

In this study, we leverage data from the three generational Young Finns Study (YFS-3G) cohort to examine the associations between a CVH-related DNAm score and early vascular changes, assessed by cIMT [Pahkala et al., 2026]. Specifically, we aim to (1) evaluate intergenerational associations between parental DNAm score and offspring cIMT, (2) assess prospective associations between DNAm score and future cIMT within individuals, and (3) examine cross-sectional relationships in two generations. By integrating epigenetic, clinical, and vascular imaging data across generations, this study seeks to advance understanding of how DNAm contributes to CVH across the life course and to evaluate its potential as a biomarker of early vascular aging.

## Methods

### Study participants

This study was conducted using data from the YFS-3G, a longitudinal, multigenerational and population-based cohort initiated in 1980 and comprising participants from the areas of five university hospitals in Finland (Turku, Tampere, Helsinki, Kuopio and Oulu) with regular follow-up in 3-to-6-year intervals until 2018-2020 [Pahkala et al., 2026]. For this analysis, we included data from two generations: the original participants (Generation 1, G1) and their parents (Generation 0, G0). DNAm and cIMT measurements were available for both generations in 2018, with additional methylation data for G1 in 2011 and cIMT follow-up in 2018. Participants with missing data on key covariates were excluded from relevant analyses.

### DNA methylation profiling, pre-processing, and normalization

For the 2011 samples, DNA was extracted from EDTA-blood collected during the YFS 2011 follow-up using the Wizard® Genomic DNA Purification Kit (Promega Corporation, Madison, WI, USA), according to the manufacturer’s instructions. DNA integrity was assessed by analyzing a subset of samples with Agilent’s Fragment Analyzer. Genome-wide DNA methylation levels were measured using Illumina Infinium MethylationEPIC BeadChips, following the manufacturer’s protocol [Mishra et al., 2023]. For the 2018 follow-up, DNA extraction was performed using the PerkinElmer Chemagic system (CMG-1074) according to the manufacturer’s instructions, with the exception that a modified Binding Buffer 2 was used for a subset of samples. Genome-wide DNA methylation profiling in the 2018 follow-up was conducted using Illumina Infinium MethylationEPIC arrays (versions 1 and 2), with data available across multiple batches in generations G0 and G1. To minimize potential batch effects and ensure consistency in the DNAm score calculation, we restricted analyses to a single array version per generation. For G0, EPIC version 2 data were selected due to the larger sample size. For G1, EPIC version 1 batch 1 was used as other batches exhibited substantial missingness in CpG sites (467/609 CpGs missing in version 1; 249/609 CpGs missing in version 2) included in the DNAm score, which could compromise score reliability.

#### Preprocessing and normalization of the 2011 methylation data

Initial quality control of samples was performed using the *minfi* package in R [Aryee et al., 2014]. Multiple criteria were applied to ensure high quality data prior to downstream analyses. First, detection p-values were assessed to evaluate probe performance within each sample. Samples were retained if the mean detection p-value across all probes was < 0.05. Samples exceeding this threshold were excluded from further analysis. Second, signal intensity distributions were examined using the *getQC* function in *minfi*. Samples were expected to cluster based on the log2 median intensities of methylated and unmethylated signals. Samples that did not cluster with the main cohort were considered outliers and removed. Third, sex prediction was performed using the *getSex* function in *minfi*. Predicted sex was compared to reported sex, and samples with discordant sex assignments were excluded. Background correction and normalisation were performed sequentially using the *preprocessNoob* and *preprocessQuantile* functions implemented in *minfi.* The *preprocessNoob* function was applied for background subtraction and dye-bias correction using the Normal-exponential Out-Of-Band (noob) method, as described by [Triche et al., 2013]. This method estimates background signal from out-of-band probes and corrects each sample individually. Dye-bias normalisation is simultaneously performed using a subset of control probes to estimate and adjust for dye-related technical variation. By default, both procedures are carried out within the function. Following background correction, stratified quantile normalisation was applied using *preprocessQuantile*. This approach performs both within and between sample normalisation. At probes level, they were retained only if they exhibited a detection p-value < 0.01 in at least 99% of the samples.

#### Preprocessing and normalization of the 2018 methylation data

The preprocessing was done using the *openSesame()* function in SeSAMe pipeline (version 1.20.0) which implements an integrated workflow including background correction, dye-bias correction, normalization, and masking of unreliable probes [Zhou et al., 2018]. Specifically, background correction was performed using the noob method, which estimates background fluorescence from out-of-band probe intensities and subtracts it on a per-sample basis. Dye-bias correction was applied to adjust for systematic differences between the red and green color channels. The pipeline further performs signal normalization to reduce technical variation while preserving biological differences. Within the SeSAMe workflow, “unreliable probes” refer to probes whose measured signal is not considered statistically distinguishable from background noise or that fail internal quality metrics. The masking of unreliable probes in SeSAMe is primarily based on detection p-values derived from signal-to-noise modeling. More specifically, SeSAMe evaluates probe performance using its pOOBAH (p-value with Out-Of-Band Array Hybridization) method. This approach estimates background signal distribution using out-of-band intensities and calculates a detection p-value for each probe in each sample. Probes with detection p-values exceeding the default threshold (p-value > 0.05) are considered not reliably detected, meaning their signal cannot be confidently distinguished from background fluorescence. These probe measurements are masked prior to downstream analysis by setting them to missing. In addition to poor detection, probes may also be masked if they exhibit extremely low signal intensity consistent with technical failure, show evidence of poor hybridization performance or map ambiguously or are prone to cross-hybridization (as defined in SeSAMe’s internal annotation resources) by masking rather than retaining these unreliable measurements, the pipeline reduces the inclusion of technical artifacts that could otherwise introduce bias or inflate false-positive findings. Only probes passing SeSAMe’s internal quality filters were retained for analysis.

### Life’s Essential 8 (LE8) framework-based DNAm score

We calculated a CVH-related DNAm score based on the LE8 framework, as previously described [Carbonneau et al., 2025]. In the original study, the DNAm score showed a moderate correlation with the corresponding phenotypic LE8 score (Pearson r=0.39 in the Framingham Heart Study and r=0.24 in the Multi-Ethnic Study of Atherosclerosis), indicating that it captures, but does not fully recapitulate measured CVH. The DNAm score was also independently associated with incident CVD and mortality, supporting its utility as a biomarker of CVH. In the present study, the DNAm score was calculated as a weighted sum of CpG-specific methylation levels using the available probes in each dataset. Of the 609 CpGs, 592 were available in G1 (2011), 533 in G1 (2018), and 412 in G0 (2018). Briefly, beta values derived from the EPIC array were combined with effect size estimates (β coefficients) obtained from the discovery EWAS. For each CpG site, methylation beta values were multiplied by their corresponding EWAS-derived effect sizes, and the DNAm score for each individual was computed as the sum of these weighted values across all included CpG sites.

### Carotid intima-media thickness (cIMT)

cIMT was assessed using high-resolution ultrasound with a 15 MHz transducer, focusing on the distal common carotid artery proximal to the bifurcation [Pahkala et al., 2026]. A 5-second ultrasound moving clip was recorded and stored for offline analysis. cIMT measurements were performed on the highest quality end-diastolic frame using a semi-automated edge-detection software (TOMTEC AutoIMT, version TTA2.41.00), with the region of interest placed 5–15 mm proximal to the carotid bifurcation. The software automatically identified the lumen–intima and media–adventitia interfaces of the far wall and calculated mean cIMT. All measurements were conducted by two blinded readers, with manual measurements performed if automated detection was inadequate. Measurements were obtained from plaque-free arterial segments.

### Traditional CVD risk factor measurements

Traditional risk factors for CVD included in this study were age, sex, body mass index (BMI), total cholesterol, high density lipoprotein (HDL) cholesterol, smoking habit, diabetes, systolic blood pressure and hypertension. The selection of the risk factors was based on the coronary risk factors used in the Framingham risk score [D’Agostino et al., 2008]. Smoking status was assessed using self-reported questionnaire data. Participants were classified based on smoking frequency into two categories: non-daily smokers (including those who never smoked or smoked less than daily) and daily smokers. Type 2 diabetes status was defined using multiple data sources, including self-reported diagnosis, use of glucose-lowering medication, biochemical measurements (fasting glucose and HbA1c levels), and, for G1 participants, register data from the Social Insurance Institution of Finland (Kela). Participants meeting any of these criteria were classified as having type 2 diabetes. Similarly, hypertension status was defined based on a combination of self-reported diagnosis, use of antihypertensive medication, and clinical measurements.

### Statistical analysis

All analyses were conducted in R (version 4.5.2). DNAm scores were standardized to have a mean of 0 and a standard deviation of 1 prior to modeling. Linear regression models were used to examine associations between DNAm scores and cIMT in four settings: intergenerational associations (G0 DNAm score in 2018 predicting G1 cIMT in 2018), prospective associations (G1 DNAm score in 2011 predicting G1 cIMT in 2018), and cross-sectional associations in both G1 and G0 (DNAm score and cIMT measured in 2018). The family clustering issue was addressed by setting the intergenerational model at offspring-level by using one parent per G1 offspring, corresponding to the n=529. Models were adjusted for age, sex, estimated cell-type proportions, and technical covariates (sentrix position), with additional adjustment for traditional cardiovascular risk factors including body mass index, systolic blood pressure, total cholesterol, HDL cholesterol, smoking status, diabetes, and hypertension. Blood cell-type proportions were estimated using reference-based DNAm deconvolution. For the 2011 dataset, cell-type proportions were estimated using the estimateCellCounts function in the *minfi* R package [Aryee et al., 2014], which implements the Houseman method [Houseman et al., 2012]. For the 2018 dataset, cell-type proportions were estimated using the *meffil* R package [Min et al., 2018].

For all analyses, three models were specified to ensure consistency across settings. Model 1 was adjusted for age, sex, estimated cell-type proportions, and technical covariates. Model 2 was additionally adjusted for traditional cardiovascular risk factors, including BMI, systolic blood pressure, total cholesterol, HDL cholesterol, smoking status, diabetes, and hypertension. Model 3 was adjusted for the same set of covariates as Model 2, with covariates defined according to the specific analytical setting. For prospective analyses, cardiovascular risk factors in Model 2 were measured at baseline (2011), whereas in Model 3 they were measured at follow-up (2018). For intergenerational analyses, Model 2 included cardiovascular risk factors measured in the offspring generation (G1), while Model 3 included the corresponding risk factors measured in the parental generation (G0). For cross-sectional analyses, Models 1 and 2 are presented, with Model 2 representing additional adjustment for contemporaneously measured cardiovascular risk factors; Model 3 was not applicable in this setting.

In addition to cIMT, association between DNAm score and carotid plaque was analyzed using the same intergenerational, prospective, and cross-sectional study designs and covariate adjustments as described for the cIMT analyses. Logistic regression models were fitted with carotid plaque presence (yes/no) as the outcome and the standardized DNAm score as the exposure.

To assess whether the prospective association between the DNAm score measured in 2011 and cIMT in 2018 reflected progression of arterial wall thickness beyond pre-existing vascular burden, we performed an additional sensitivity analysis including cIMT measured in 2007 as an additional covariate. The same covariate structures as in Models 2 and 3 were applied, with 2007 cIMT included as a measure of baseline arterial thickness.

### Genome-wide association analysis of the DNAm score

We conducted a genome-wide association study (GWAS) of the DNAm score using PLINK 2.0. Genomic DNA from the YFS participants was extracted from peripheral blood leukocytes using a commercially available kit and Qiagen BioRobot M48 Workstation according to the manufacturer’s instructions (Qiagen, Hilden, Germany). Genotyping was done using custom build Illumina Human 670 k BeadChip atWelcome Trust Sanger Institute. Genotypes were called using Illuminus clustering algorithm. Samples that failed Sanger genotyping pipeline quality control criteria (i.e., duplicated samples, heterozygosity, low call rate, or Sequenom fingerprint discrepancy) were excluded from the analysis. Similarly, samples with sex discrepancy, low genotyping call rate (<0.95) and possible relatedness (pi-hat>0.2) were excluded from the analysis. Single Nucleotide Polymorphisms (SNPs) were excluded based on Hardy-Weinberg equilibrium test (p-value≤1 × 10^−06^), failed missingness test (call rate<0.95) and failed frequency test (minor allele frequency<0.01). Overall, 546677 genotyped SNPs passed the quality control. Genotype imputation was performed on the TOPMed imputation server using TOPMed r1 reference panel [Taliun et al., 2021]. Association testing was performed using linear regression under an additive genetic model, adjusting for age, sex and the first 5 genetic principal components. Given the modest sample size (∼1300 individuals), we interpreted the results as exploratory. Genome-wide significance was defined as p-value < 5×10⁻⁸, and suggestive associations were defined as p-value < 1×10⁻⁵.

### Transcriptome-wide association and pathway enrichment analysis

We performed a transcriptome-wide association analysis to identify transcriptomic correlates of the DNAm score using gene expression levels as outcome. For each gene, we fitted a linear regression model to estimate the association between expression level and the DNAm score, adjusting for age, sex, estimated cell-type proportions, and technical covariates (the first five principal components derived from DNAm control probes). This analysis was conducted using data from the 2011 follow-up, as both transcriptomic and DNAm data were available only at this time point. Whole-blood transcriptomic profiling was performed as previously described [Mishra et al., 2022]. Briefly, gene expression was measured using the Illumina HumanHT-12 v4 Expression BeadChip, and standard quality control, background correction, quantile normalization and log2 transformation was applied. Genes with false discovery rate (FDR) < 0.05 were considered statistically significant. To investigate the biological relevance of DNAm score-associated genes, Gene Ontology Biological Process enrichment analysis was performed using *clusterProfiler* R package [Wu et al., 2021].

## Results

### Study population characteristics

A total of 1,432 YFS-3G participants contributed to at least one analysis, comprising the original cohort (G1; mean age 47 ± 5 years, 53.9% female) and their parents (G0; mean age 73 ± 5 years, 51.8% female). Analytical sample sizes varied by study design and data availability, as described below and shown in Table 1 and Supplementary Tables S1–S2. Study population characteristics of the intergenerational analysis cohort are summarized in Table 1. In general, G0 participants exhibited a higher prevalence of hypertension and diabetes, whereas G1 participants had lower cIMT values and more favorable lipid profiles. The mean DNAm score was higher in G1 than in G0, indicating better epigenetic CVH, (0.006 vs. 0.003, p-value < 0.001), consistent with the younger age and lower risk factor burden in G1.

**Table 1.** Population characteristics of the intergenerational analysis cohort (G0 parents and G1 offspring). Data are presented as mean ± standard deviation (SD), median [interquartile range (IQR)] or n (%). Percentage were calculated using the number of participants with no-missing data for each variable; therefore, denominators may differ from the total number of participants. DNAm score values are presented on the original scale. Standardized values were used only for regression analyses.

| Characteristic | G1 (Original cohort) | G0 (Parents of G1) |
| --- | --- | --- |
| Number of participants | 529 | 903 |
| Age, years (mean ± SD) | 47 ± 5 | 73 ± 5 |
| Female, n (%) | 285 (53.9) | 468 (51.8) |
| Body Mass Index (BMI), kg/m<sup>2</sup> (mean ± SD) | 27.7 ± 5.2 | 28.5 ± 5.1 |
| Waist circumference, cm (mean ± SD) | 94.7 ± 14.2 | 99.5 ± 14.0 |
| Systolic blood pressure, mmHg (mean ± SD) | 128 ± 15 | 142 ± 21 |
| Diastolic blood pressure, mmHg (mean ± SD) | 82 ± 10 | 79 ± 10 |
| Hypertension, n (%) | 74 (14.1) | 485 (55.5) |
| Total cholesterol, mmol/L (mean ± SD) | 5.1 ± 0.9 | 4.9 ± 1.1 |
| HDL cholesterol, mmol/L (mean ± SD) | 1.3 ± 0.4 | 1.3 ± 0.4 |
| LDL cholesterol, mmol/L (mean ± SD) | 3.1 ± 0.8 | 2.9 ± 1.0 |
| Triglycerides, mmol/L (mean ± SD) | 1.5 ± 1.1 | 1.5 ± 0.8 |
| Diabetes, n (%) | 37 (7.0) | 310 (34.6) |
| Smoking status, n (%) | 69 (13.2) | 50 (6.0) |
| Alcohol consumption, standard drinks/day (mean ± SD) | 0.68 ± 0.95 | 0.59 ± 1.0 |
| Physical activity score (MET hour/week) | 19.3 ± 17.5 | 16.7 ± 18.0 |
| C-Reactive Protein, mg/L (median [IQR]) | 1.2 [0.6-2.5] | 1.3 [0.7-2.7] |
| DNAm Score (mean ± SD) | 0.006 ± 0.002 | 0.002 ± 0.002 |
| Common carotid intima media thickness (cIMT), mm (mean ± SD) | 0.61 ± 0.12 | 0.83 ± 0.19 |

### Intergenerational association between parental DNAm score and offspring cIMT

Parents with higher DNAm scores had offspring with lower cIMT. In age- and sex-adjusted linear models (Model 1; with additional control for parents’ leukocyte proportions and technical covariates), a 1 SD higher parental (G0) DNAm score was associated with a 0.02 mm lower offspring (G1) cIMT (*β* = –0.022 per SD, 95% CI [–0.040, –0.004], p-value = 0.02). This association remained significant after further adjusting for the offspring’s own cardiovascular risk factors (BMI, systolic blood pressure, total and HDL cholesterol, smoking, diabetes, and hypertension status in 2018) (Model 2). However, when additionally adjusting for parental (G0) cardiovascular risk factors (Model 3), the association was attenuated and no longer statistically significant (*β* = –0.030 per SD, 95% CI [–0.066, 0.002], p-value = 0.07). These results, summarized in Table 2, suggest a potential intergenerational association between parental DNAm profiles and offspring vascular health; however, the attenuation after adjustment for parental risk factors indicates that this relationship may be partly explained by shared familial or parental characteristics and should therefore be interpreted with caution.

**Table 2.** Association between the DNAm score and carotid intima-media thickness (cIMT) across intergenerational and life-course models. Model 1 was adjusted for age, sex, estimated cell-type proportions, and technical covariates. Model 2 was additionally adjusted for traditional cardiovascular risk factors, including body mass index, systolic blood pressure, total cholesterol, HDL cholesterol, smoking status, diabetes, and hypertension. Model 3 included the same covariates as Model 2, with definitions tailored to the analytical setting: in prospective analyses, risk factors were measured at baseline (2011) for Model 2 and at follow-up (2018) for Model 3; in intergenerational analyses, Model 2 included offspring (G1) risk factors, whereas Model 3 included parental (G0) risk factors. For cross-sectional analyses, only Models 1 and 2 are presented, with Model 2 reflecting adjustment for contemporaneously measured cardiovascular risk factors (Model 3 not applicable).

| Analysis type | Sample size (n) | Model 1 |  |  | Model 2 |  |  | Model 3 |  |  |
| --- | --- | --- | --- | --- | --- | --- | --- | --- | --- | --- |
| | | Beta ($\beta$) per 1 SD DNAm Score | 95% CI | P-value | Beta ($\beta$) per 1 SD DNAm Score | 95% CI | P-value | Beta ($\beta$) per 1 SD DNAm Score | 95% CI | P-value |
| Intergenerational (G0 to G1) | 529 | -0.022 | -0.040 to -0.004 | 0.018 | -0.019 | -0.038 to -0.0006 | 0.04 | -0.03 | -0.07 to 0.002 | 0.07 |
| Prospective (G1, 2011 to 2018) | 1246 | -0.030 | -0.044 to -0.017 | 1.1 x 10<sup>-5</sup> | -0.014 | -0.029 to 0.002 | 0.08 | -0.019 | -0.034 to -0.005 | 0.009 |
| Cross-sectional (G1, 2018) | 249 | -0.037 | -0.069 to -0.005 | 0.026 | -0.029 | -0.065 to -0.007 | 0.113 | - | - | - |
| Cross-sectional (G0, 2018) | 903 | -0.02 | -0.05 to -0.0009 | 0.04 | -0.027 | -0.052 to -0.002 | 0.034 | - | - | - |

### Prospective association between baseline DNAm score and future cIMT

A higher DNAm score in G1 was prospectively associated with lower cIMT measured seven years later. In the minimally adjusted model (Model 1), controlling for age, sex, cell composition, and technical factors, the 2011 DNAm score was a significant predictor of 2018 cIMT (*β* = –0.030 per SD, 95% CI [–0.044, –0.017], p-value = 1.1×10^−5^). This corresponds to approximately a 0.03 mm thinner carotid intima-media per SD higher baseline DNAm score. After additional adjustment for baseline (2011) cardiovascular risk factors (Model 2), the association was attenuated and no longer statistically significant (*β* = –0.014 per SD, 95% CI [–0.029, 0.002], p-value = 0.08). In contrast, when adjusting for risk factors measured at follow-up (2018; Model 3), the association was stronger and statistically significant (*β* = –0.019 per SD, 95% CI [–0.034, −0.004], p-value = 0.009). These findings indicate that the prospective association between DNAm score and cIMT is sensitive to the choice and timing of covariate adjustment. While the minimally adjusted model suggests a robust inverse association, adjustment for baseline risk factors attenuates the effect, whereas adjustment for follow-up covariates yields a stronger association, the interpretation of which should be made with caution given the potential for overadjustment or conditioning on variables measured contemporaneously with the outcome.

In a sensitivity analysis additionally adjusting for cIMT measured in 2007, the association between the 2011 DNAm score and cIMT in 2018 was further attenuated and no longer statistically significant in either Model 2 (p-value=0.46) or Model 3 (p-value=0.26). These findings indicate that although the DNAm score is associated with later cIMT burden, the association is largely explained by pre-existing arterial wall thickness. Thus, the DNAm score appears to reflect cumulative CVH and vascular burden rather than predicting subsequent cIMT progression independently of prior cIMT.

### Cross-sectional associations between DNAm score and cIMT in G0 and G1

In cross-sectional analyses of the 2018 examination, the DNAm score was inversely associated with cIMT in both generations. In G1, a 1 SD higher DNAm score was associated with 0.037 mm lower cIMT (*β* = –0.037, 95% CI [–0.069, –0.005], p*-*value = 0.03) after adjusting for age, sex, cell-type proportions, and technical covariates. A similar association was observed in G0 (parents) in 2018 (*β* = –0.022, 95% CI [–0.040, –0.004], p-value = 0.02). However, while the inclusion of the traditional cardiovascular risk factors in these cross-sectional models caused attenuation of association in G1 (fully adjusted p-value = 0.09), in G0, the association remained significant with similar magnitude (fully adjusted p-value = 0.03). These results imply that the DNAm score’s cross-sectional relationship with cIMT is partly explained by its overlap with concurrent risk factors (e.g., older individuals and those with higher blood pressure or adverse lipids tend to have both lower DNAm scores and higher cIMT). Nevertheless, the consistent direction of effect across generations supports the association between higher DNAm scores (better epigenetic CVH) and lower cIMT. Although the DNAm score was derived from LE8 metrics, we did not observe strong or consistent cross-sectional associations between the DNAm score and traditional cardiovascular risk factors in either generation. In fully adjusted models, associations with BMI, total and HDL cholesterol, smoking, and systolic blood pressure were weak and not statistically significant [Table 3]. The positive cross-sectional associations of age, male sex and smoking with the DNAm score should not be interpreted as indicating that these characteristics are beneficial for CVH. Rather the DNAm score is a weighted composite derived from EWAS of LE8 traits and may capture cumulative methylation patterns influenced by multiple biological and exposure-related processes, not one-to-one effects of individual risk factors. This interpretation is supported by the score’s robust prospective and intergenerational associations with cIMT, despite its modest cross-sectional correlations with conventional risk factors.

**Table 3.** Cross-sectional associations between the DNAm score and traditional cardiovascular risk factors in G0 and G1. Models are adjusted for age, sex, leukocyte cell-type proportions and technical covariates. Beta (β) coefficients represent the association between each risk factor and the DNAm score.

| Risk factor | G0 (parents) $\beta$ (95% CI) | P-value | G1 (original cohort) $\beta$ (95% CI) | P-value |
| --- | --- | --- | --- | --- |
| Age | 0.008 (0.006 to 0.011) | $2.2 \times 10^{-10}$ | 0.005 (0.0000 to 0.0105) | 0.05 |
| Sex, male | 0.055 (0.026 to 0.083) | 0.0002 | 0.066 (0.025 to 0.107) | 0.002 |
| Body Mass Index (BMI) | -0.06 (-0.117 to -0.003) | 0.039 | 0.026 (-0.1007 to 0.152) | 0.691 |
| Smoking (current vs. not) | 0.061 (0.007 to 0.115) | 0.027 | -0.01 (-0.064 to 0.043) | 0.694 |
| Total Cholesterol | 0.013 (-0.0003 to 0.0255) | 0.055 | -0.006 (-0.024 to 0.012) | 0.511 |
| HDL Cholesterol | -0.016 (-0.055 to 0.024) | 0.438 | 0.009 (-0.044 to 0.062) | 0.73 |
| Systolic Blood Pressure | 0.0005 (-0.0001 to 0.0012) | 0.11 | 0.0011 (-0.0000 to 0.0023) | 0.055 |
| Diabetes (yes vs. no) | 0.015 (-0.014 to 0.045) | 0.32 | 0.0143 (-0.053 to 0.082) | 0.68 |
| Hypertension (yes vs. no) | 0.003 (-0.024 to 0.03) | 0.849 | 0.002 (-0.046 to 0.05) | 0.94 |

### Association between the DNAm score and carotid plaque

We next examined whether the DNAm score was associated with carotid plaque, a marker that more directly reflects focal atherosclerotic lesion development than cIMT. Overall, evidence for an association between the DNAm score and carotid plaque was limited. In the intergenerational analysis, parental DNAm score was not associated with offspring plaque status in any model (all p-values > 0.05) [Supplementary Table S5]. Similarly, no significant cross-sectional associations were observed in either G1 or G0 after adjustment for covariates. In the prospective analysis, a higher DNAm score measured in 2011 was associated with lower odds of carotid plaque in 2018 in the minimally adjusted model (log-odds per SD = −0.25, 95% CI −0.50 to −0.006, p-value = 0.045). However, this association was attenuated after adjustment for traditional cardiovascular risk factors and was no longer statistically significant in Models 2 and 3. Taken together, these findings suggest that the DNAm score shows a stronger and more consistent relationship with cIMT than with carotid plaque and that any association with plaque burden may be largely explained by established cardiovascular risk factors.

### Transcriptomic signatures associated with the DNAm score

In the transcriptome-wide association study (TWAS), the DNAm score was associated with expression levels of 3275 genes at FDR < 0.05. Gene Ontology Biological Process enrichment analysis identified 208 enriched biological processes. Overall, the enriched biological processes were predominantly related to inflammatory and immune processes, indicating that variation in the DNAm score is accompanied by broad differences in immune and inflammation related transcriptomic activity [Figure 1]. These findings suggest that the DNAm score captures molecular signatures linked to systemic immune regulation, considtent with the broader role of inflammation in vascular aging and atherosclerosis.

**Figure 1.**
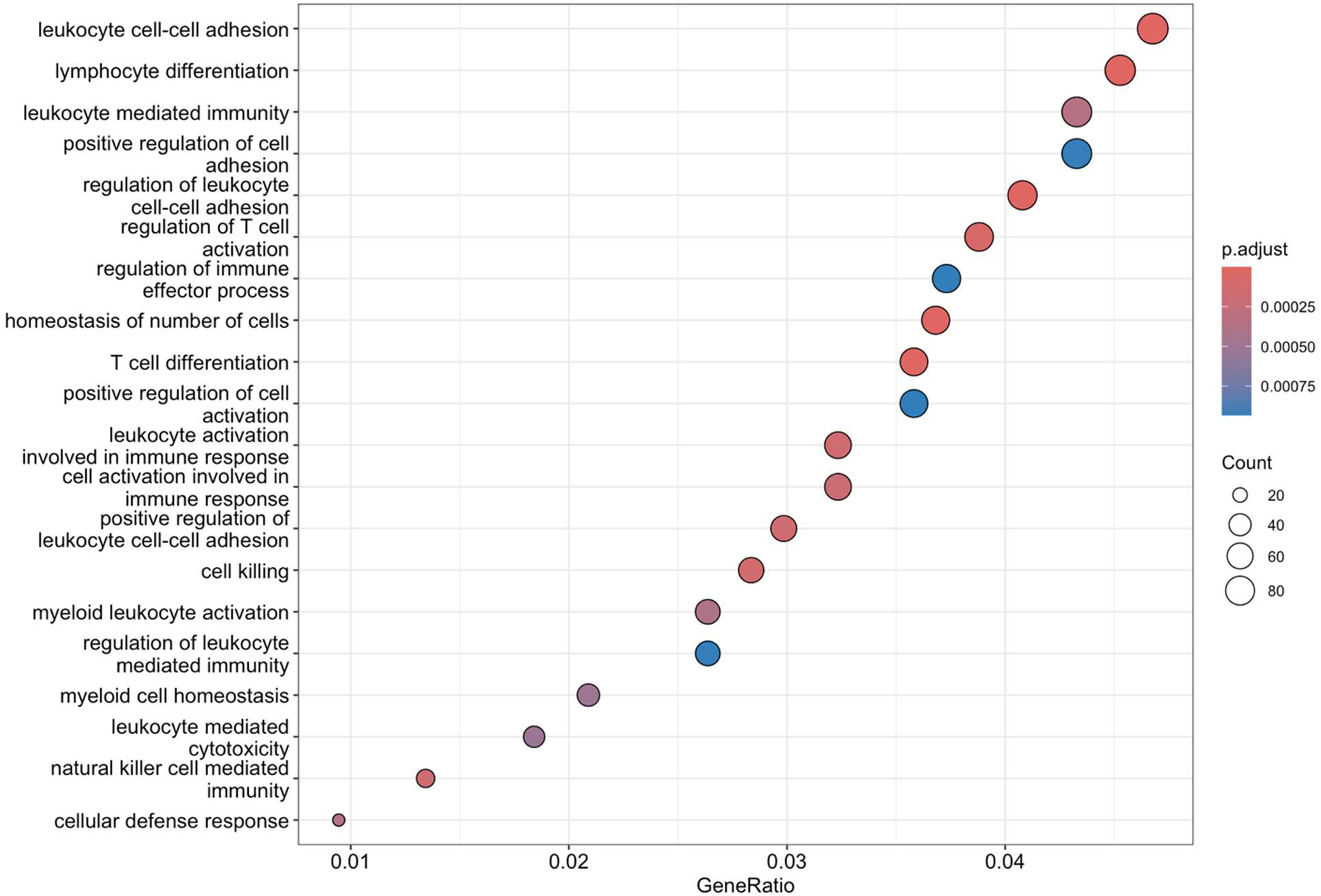
The top 20 enriched Gene Ontology (GO) Biological Process enrichment of genes associated with the DNAm score. Each dot represents a GO term, with dot size corresponding to the number of genes contributing to the pathway and dot color indicating the significance of enrichment (adjusted p-value using the Benjamini–Hochberg method). The x-axis shows the gene ratio, defined as the proportion of input genes assigned to each GO term.

### Genetic determinants of the DNAm score: a genome-wide association analysis

In the GWAS of the DNAm score (N = 1320), the genomic inflation factor was λ = 0.99, indicating well calibrated test statistics [Supplementary Figures S1 and S2]. Two variants reached genome-wide significance (p-value < 5×10⁻⁸). The most significant variant was located on chromosome 9 at position 85,351,004 (chr9:85351004; β = −0.00476, SE = 0.00086, p-value = 3.65 × 10⁻⁸), and the second on chromosome 4 at position 32,006,132 (chr4:32006132; β = −0.00463, SE = 0.00084, p-value = 4.70 × 10⁻⁸). In addition, 258 variants showed suggestive associations (p-value < 1×10⁻⁵), including a cluster of SNPs on chromosome 4 (positions ∼32.1–32.3 Mb) with consistent direction of effect and similar allele frequencies, suggesting a potential regional signal. The SNPs were further analyzed using SnpXplorer v2 [Tesi et al., 2021]. Of the nominally significant variants, 13 have been previously reported in GWAS of related traits: seven in LDL cholesterol, three in telomere length, two in eosinophil counts, and one in ocular traits [Figure 2] [Supplementary Table S4]. These overlaps support the biological relevance of the DNAm score and suggest it may reflect genetic influences linked to lipid metabolism, cellular aging, immune function, and systemic health. Full results are provided in Supplementary Table S4.

**Figure 2.**
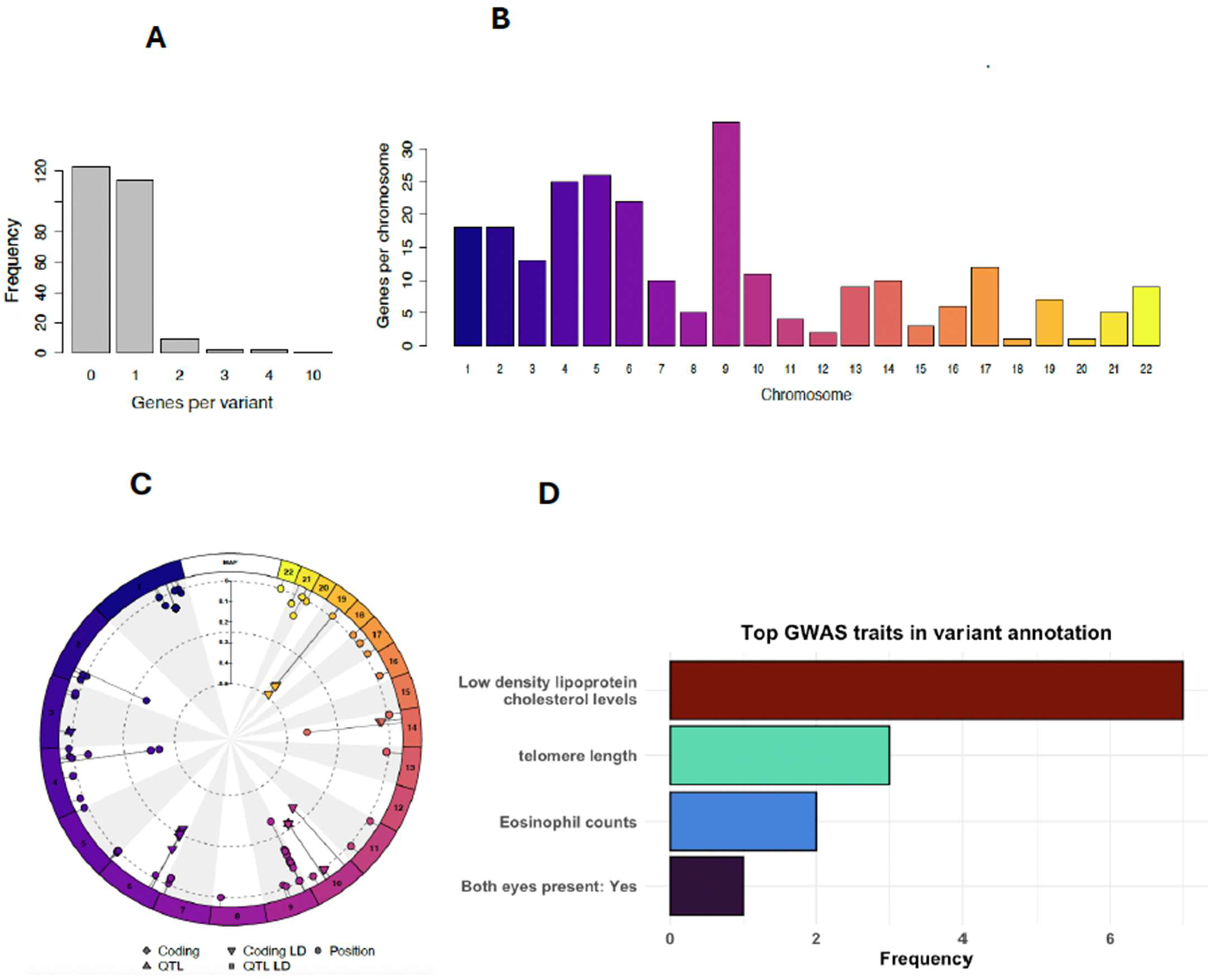
Results of the functional annotation of the 260 SNPs (single nucleotide polymorphisms) associated with DNA methylation (DNAm) score. (**A**) Number of genes associated with each of the 260 SNPs. (**B**) Chromosomal distribution of all the 260 SNPs. (**C**) Circular summary figure showing the type of annotation of each SNP (coding, eQTL or annotated by their positions) as well as each SNP’s minor allele frequency and chromosomal distribution. (**D**) Number of genes associated with the 260 independent SNPs, expressed as frequency, for which a previous association was reported in the OpenGWAS (https://opengwas.io/).

## Discussion

In this multigenerational cohort study, we investigated intergenerational, prospective and cross-sectional associations between a CVH-related DNAm score and early vascular changes, as measured by cIMT. Our findings provide novel evidence that epigenetic signatures of CVH are associated with vascular structure across the life course and across generations.

The most notable finding is the observed intergenerational association between parental DNAm score and offspring cIMT. Higher parental DNAm scores were associated with lower cIMT in their adult children in models adjusted for age, sex, and technical factors, and this association persisted after accounting for the offspring’s own cardiovascular risk factors. This pattern suggests that DNAm profiles may capture familial influences on CVH that extend beyond the offspring’s measured risk factor profile. However, the attenuation of the association after additional adjustment for parental cardiovascular risk factors indicates that this relationship is at least partly explained by shared familial or parental characteristics. This finding tempers the interpretation of a direct intergenerational epigenetic effect and instead points toward a combination of mechanisms, including shared environment, behavioral transmission, and correlated cardiometabolic risk profiles within families. While prior studies have demonstrated familial aggregation of cardiovascular risk and subclinical atherosclerosis [Sacco et al., 2009; Lie et al., 2019], our results extend this literature by suggesting that DNAm-based CVH scores may reflect these intergenerational patterns. However, the lack of robustness to adjustment for parental risk factors underscores the need for cautious interpretation and further work to disentangle epigenetic contributions from shared familial confounding.

The prospective association observed in the original cohort supports the potential biological relevance of the DNAm score in early vascular changes. A higher DNAm score was associated with lower cIMT measured seven years later in minimally adjusted models, suggesting that DNAm profiles may capture processes related to the early development of atherosclerosis. This is consistent with previous work indicating that DNAm integrates cumulative effects of lifestyle, metabolic, and inflammatory exposures over time [Westerman et al., 2020; Zheng et al., 2022; Yousefi et al., 2022]. However, the association was attenuated and no longer statistically significant after adjustment for baseline cardiovascular risk factors, indicating that part of the DNAm signal overlaps with established risk factors measured at the same time point. In contrast, the association remained statistically significant when adjusting for contemporaneous risk factors measured at follow-up. This pattern should be interpreted with caution, as adjustment for covariates measured contemporaneously with the outcome may introduce overadjustment or collider bias, particularly if these variables lie on the causal pathway between baseline DNAm and subsequent vascular changes. Taken together, these findings suggest that while the DNAm score is associated with future cIMT, its independent predictive value beyond traditional cardiovascular risk factors remains uncertain. Rather than acting as a fully independent predictor, the DNAm score may capture cumulative and potentially upstream biological variation linked to cardiovascular risk, complementing rather than replacing conventional risk factor assessment.

An important additional observation was that the prospective association between the DNAm score and cIMT was no longer statistically significant after further adjustment for cIMT measured four years before DNAm assessment. This suggests that the DNAm score is associated with later vascular burden but does not appear to predict cIMT independently of pre-existing arterial thickness. In other words, the DNAm score may primarily capture the cumulative biological consequences of prior cardiometabolic exposures that are already reflected in arterial wall structure rather than providing independent information on subsequent cIMT progression. This interpretation is consistent with the view that DNAm-based CVH scores function as integrative biomarkers of long-term CVH rather than direct predictors of vascular disease progression.

In cross-sectional analyses, the DNAm score was inversely associated with cIMT in both generations, although the association was attenuated after adjustment for traditional risk factors, particularly in the younger generation. This pattern suggests that cross-sectional relationships may be more influenced by concurrent exposures and risk factor burden, whereas longitudinal associations better reflect the cumulative and temporally stable nature of epigenetic signatures.

In contrast to the cIMT findings, we observed little evidence that the DNAm score was associated with carotid plaque. The only nominally significant association was observed in the prospective analysis, where a higher baseline DNAm score was associated with lower odds of plaque at follow-up in the minimally adjusted model. However, this association was attenuated after adjustment for traditional cardiovascular risk factors. The discrepancy between the cIMT and plaque results may reflect differences in the biological processes captured by these vascular phenotypes. While carotid plaque is generally considered a more specific marker of atherosclerosis, cIMT reflects broader arterial wall thickening arising from multiple processes, including ageing, blood pressure-related medial hypertrophy, adaptive vascular remodeling, and early atherosclerotic change. The stronger association with cIMT therefore suggests that the DNAm score may primarily capture cumulative biological processes related to vascular ageing and arterial remodeling rather than focal plaque formation. Nevertheless, given the established role of inflammation in atherosclerotic plaque development, larger studies will be required to determine whether DNAm-based CVH scores are associated with plaque burden or progression.

Our findings are consistent with emerging literature demonstrating that DNAm-based scores derived from CVH metrics are associated with cardiovascular outcomes and risk factors [Zheng et al., 2022; Carbonneau et al., 2025]. For example, prior epigenome-wide association studies have identified methylation signatures linked to smoking [Mishra et al., 2020], lipid metabolism [Gomez-Alonso et al., 2021], and inflammation [Ligthart et al., 2016], all of which contribute to atherosclerosis development. The DNAm score used in this study, based on the LE8 framework, builds on this work by integrating multiple CpG sites into a composite measure of CVH.

The TWAS and Gene Ontology enrichment analyses provide additional biological context for the observed associations between the DNAm score and cIMT. The predominance of immune- and inflammation-related biological processes among DNAm score-associated genes suggests that this methylation score may reflect systemic inflammatory and immune activity relevant to early atherosclerotic changes. This interpretation is consistent with the established role of inflammation in atherosclerosis and with the concept that blood-based DNAm signatures capture cumulative biological responses to cardiometabolic and environmental exposures. However, it is important to note that cIMT is a continuous marker of arterial wall thickening and is not specific to atherosclerosis. Increased cIMT may also reflect ageing and hypertension-related medial hypertrophy or adaptive vascular remodeling, whereas atherosclerosis per se is more directly characterized by lipid-rich inflammatory plaque formation, better captured by measures such as carotid plaque. Accordingly, because these analyses are observational and based on whole-blood molecular profiles, the findings should be interpreted as supporting biological plausibility rather than demonstrating a causal pathway linking the DNAm score, gene expression, and vascular structure.

Importantly, earlier studies have shown that DNAm markers can predict incident cardiovascular disease and mortality [Zheng et al., 2022; Carbonneau et al., 2025], but fewer have examined subclinical vascular phenotypes such as cIMT or explored life-course and intergenerational dimensions. Our results therefore add to the literature by demonstrating that DNAm scores are associated not only with clinical endpoints but also with early vascular changes that precede clinical disease outcomes.

The relatively weak cross-sectional associations between the DNAm score and individual traditional risk factors in our study are also noteworthy. This aligns with previous observations that DNAm markers often reflect complex, cumulative biological processes rather than single risk factor pathways [Westerman et al., 2020; Zheng et al., 2022; Yousefi et al., 2022]. Thus, DNAm scores may serve as integrative biomarkers that capture latent disease risk beyond what is measurable through conventional clinical variables at a single time point.

Several mechanisms may explain the observed associations. DNAm is known to regulate gene expression [Jones PA. 2012] and is responsive to environmental exposures such as diet, smoking, physical activity, and psychosocial stress [Fraga et al., 2005; Gluckman et al., 2009; Baccarelli et al., 2012]. These exposures influence key pathways involved in atherosclerosis, including inflammation, endothelial dysfunction, and lipid metabolism.

The intergenerational findings may reflect a combination of shared genetics, shared environment, and potential epigenetic inheritance mechanisms. While most DNAm patterns are reprogrammed during gametogenesis, some evidence suggests that environmentally induced epigenetic marks may persist across generations or influence offspring phenotype indirectly through parental exposures [Gluckman et al., 2007; Fitz-James et al., 2022; Klibaner-Schiff et al., 2024]. Additionally, parental DNAm profiles may act as proxies for long-term family-level environmental and behavioral patterns that shape offspring CVH.

To evaluate whether the association between the DNAm score and cIMT could be attributed to underlying genetic variation, we conducted a GWAS of the DNAm score in 1,320 individuals. Two SNPs reached genome-wide significance (p-value < 5×10⁻⁸), and several additional variants showed suggestive associations (p-value < 1×10⁻⁵). Notably, 13 of these nominally significant SNPs have been previously reported in GWAS of related traits, including seven in LDL cholesterol, three in telomere length, two in eosinophil counts, and one in a GWAS of ocular traits (both eyes present). These overlaps suggest that the DNAm score may capture genetic influences relevant to lipid metabolism, cellular aging, immune function, and systemic health. This convergence with established GWAS findings supports the biological plausibility of the DNAm score and highlights its potential as an integrative biomarker reflecting diverse pathways implicated in CVH. These findings also suggest that the DNAm score is not predominantly driven by common genetic variation and may instead reflect environmental or non-genetic biological processes. This supports the interpretation that the DNAm score captures cumulative exposures relevant to early vascular changes, rather than merely indexing inherited genetic risk. However, given the modest sample size, these findings should be interpreted cautiously and warrant replication in larger cohorts.

Several limitations should be acknowledged. First, the observational design precludes causal inference. Second, DNAm was measured in whole blood, which reflect circulating CVD risk factor status rather than direct methylation patterns in vascular tissues. Third, residual confounding by unmeasured environmental or genetic factors cannot be excluded. Finally, the generalizability of findings may be limited to populations of European ancestry.

In conclusion, our study extends the current knowledge by showing that a CVH-related DNAm score, based on the LE8 framework, is also associated with early vascular changes across the life course and, to some extent, across generations. While consistent inverse associations with cIMT were observed in prospective, cross-sectional, and intergenerational analyses, these relationships were sensitive to adjustment for traditional cardiovascular risk factors, indicating partial overlap with established traditional risk factor pathways. These findings suggest that DNAm captures biologically meaningful information related to CVH, likely reflecting cumulative and integrated effects of cardiometabolic exposures over time. However, the attenuation of associations after risk factor adjustment indicate that the DNAm score should not be interpreted as an independent predictor of vascular outcomes. Instead, DNAm-based scores may serve as complementary holistic integrative biomarkers that provide insight into early vascular aging and the biological embedding of cardiovascular risk. Future studies are needed to clarify the causal pathways underlying these associations, disentangle epigenetic effects from shared familial and environmental influences, and determine whether incorporating DNAm markers into risk assessment frameworks can meaningfully improve early prevention strategies.

## Data Availability

The dataset supporting the conclusions of this article were obtained from the Cardiovascular Risk in Young Finns study which comprises health related participant data. The use of data is restricted under the regulations on professional secrecy (Act on the Openness of Government Activities, 612/1999) and on sensitive personal data (Personal Data Act, 523/1999, implementing the EU data protection directive 95/46/EC). Due to these restrictions, the data cannot be stored in public repositories or otherwise made publicly available. Data access may be permitted on a case-by-case basis upon request only. Data sharing outside the group is done in collaboration with YFS group and requires a data-sharing agreement. Investigators can submit an expression of interest to the chairman of the publication committee, Prof Olli Raitakari (University of Turku, Finland), Prof Mika Kähönen (Tampere University, Finland) and Prof Terho Lehtimäki (Tampere University, Finland). Requests to access these datasets should be directed to OR; TL; MK.

